# Scoping review of knowledge graph applications in biomedical and healthcare sciences

**DOI:** 10.1101/2023.12.13.23299844

**Authors:** Sanjay Budhdeo, Joe Zhang, Yusuf Abdulle, Paul M Agapow, Douglas GJ McKechnie, Matt Archer, Viraj Shah, Eugenia Forte, Ayush Noori, Marinka Zitnik, Hutan Ashrafian, Nikhil Sharma

## Abstract

**Introduction:** There is increasing use of knowledge graphs within medicine and healthcare, but a comprehensive survey of their applications in biomedical and healthcare sciences is lacking. Our primary aim is to systematically describe knowledge graph use cases, data characteristics, and research attributes in the academic literature. Our secondary objective is to assess the extent of real-world validation of findings from knowledge graph analysis.

**Methods:** We conducted this review in accordance with the PRISMA extension for Scoping Reviews to characterize biomedical and healthcare uses of knowledge graphs. Using keyword-based searches, relevant publications and preprints were identified from MEDLINE, EMBASE, medRxiv, arXiv, and bioRxiv databases. A final set of 255 articles were included in the analysis.

**Results:** Although medical science insights and drug repurposing are the most common uses, there is a broad range of knowledge graph use cases. General graphs are more common than graphs specific to disease areas. Knowledge graphs are heterogenous in size with median node numbers 46 983 (IQR 6 415-460 948) and median edge numbers 906 737 (IQR 66 272-9 894 909). DrugBank is the most frequently used data source, cited in 46 manuscripts. Analysing node and edge classes within the graphs suggests delineation into two broad groups: biomedical and clinical. Querying is the most common analytic technique in the literature; however, more advanced machine learning techniques are often used.

**Discussion:** The variation in use case and disease area focus identifies areas of opportunity for knowledge graphs. There is diversity of graph construction and validation methods. Translation of knowledge graphs into clinical practice remains a challenge. Critically assessing the success of deploying insights derived from graphs will help determine the best practice in this area.

## Introduction

### Context and importance of knowledge graphs

Data representation plays a vital role in advancing medicine: efficient organisation of increasingly large datasets enables analysis to robustly generate novel insights. (1) (2) An increasingly important representation method is the knowledge graph (KG). KGs consist of the entities, relationships, and facts in a given domain, captured as a graph of nodes (representing entities or concepts) and edges (indicating the relationships or associations between them), often enriched with attributes, classifications, and semantic meanings. (3)

Data representation through KGs has a historical foundation dating back multiple decades. Key accelerants to adoption were the development of Resource Description Framework (RDF) and Web Ontology Language (OWL) standards for the semantic web and Google’s implementation of knowledge graphs in its search algorithms. (3) (4) KGs offered distinct advantages over traditional relational databases, including greater schema flexibility and the capacity to capture nuanced capture of edge characteristics and relationships, making them valuable for knowledge retrieval analytical purposes such as recommender systems. (3) Early uses of KGs within healthcare included theoretical conceptualizations of clinical reasoning (5) and in research literature representation. (6) More recent applications in biomedicine and healthcare have included identifying drug repurposing candidates and generating novel biomedical hypotheses by established and early-stage pharmaceutical companies such as AstraZeneca (7) and Benevolent AI. (8)

### Motivation for review

Given the increasing use of KGs in biomedical and healthcare sciences, this review presents a comprehensive survey of research literature to detail the use of KGs within this domain. Many other reviews discuss the history of KGs and the methodologies used in their construction and analysis. There are also reviews that categorize uses for knowledge graphs. (3) This prior work includes systematic overviews of specific methods or analysis archetypes for KGs, for example, KG reasoning, (9) KG completion, (10) and relational machine learning. (11) There have also been reviews on data quality and methodology in KG construction across uses, including KG completeness (12) and generative KG construction. (13)

There are commentaries and reviews for specific use cases within biomedicine, such as drug repurposing and adverse drug reactions (14), and focussed reviews, for example, on bioinformatic graphs and their analyses without a systematic survey of use cases. (15) In addition, some reviews have surveyed use of KGs in specific disease areas. (16) There are also commentary articles that outline potential broader use cases for biomedical KGs. (17)

Despite this work, no systematic survey of use cases for knowledge graphs within the biomedical and healthcare sciences has been conducted. Our primary aim is to address this gap by providing a landscaping of disease areas in which knowledge graphs are used, and how they are used. We survey characteristics of these manuscripts, including author affiliations and funding. We also review graph characteristics: including node class and edge class numbers, node and edge numbers, node class domains and whether graphs have been made openly available. We review techniques used in KG analysis.

Our secondary research question examines the extent to which findings or insights from knowledge graphs been tested and validated in the real-world. There has been no systematic survey KG validation in the biomedical literature. KG validation includes the process of ensuring the accuracy of the data within a KG, as well as verifying insights derived from the KG. Validation is often divided into two categories: internal and external; however, these terms do not have standardized definitions. Consequently, in this article, we use the terms ’inside graph validation’ and ’outside graph validation’. Inside graph validation assesses robustness of the graph based on the graph data itself. This includes hold-one-out studies or cross-validation, calculation and assessment of performance scores, or analysis with multiple algorithms on the same graph and comparing results. Outside graph validation refers to testing of insights in a different dataset, for example through in vitro testing or a clinical trial.

### Aims of the review

The primary aim of this review is to establish a systematic description of the use cases, data characteristics, and research characteristics in current KG implementations in the academic literature. The secondary aim is to determine the extent to which findings or insights from KGs have been tested and validated in the real world.

## Methods

### Protocol

We were guided by the PRISMA extension for Scoping Reviews checklist. (18) A protocol was written *a priori* and posted on the website OSF on 5 November 2021 (https://osf.io/etg6f). We limited our search to only academic literature (i.e., publications and preprints), and did not survey other sources of insights such as patent databases. This topic is suitable for a scoping review as it represents an exploratory mapping of the literature, where it was not possible to delineate how best to categorise manuscripts *a priori*.

### Literature Search

Relevant studies were be identified from an academic search on databases for biomedical literature. We used MEDLINE and EMBASE (via Ovid), as well as medRxiv, arXiv (section: quantitative biology) and bioRxiv, employing free-text searches for keywords. Our search strategy was as follows: “knowledge graph” OR “graph neural network” OR “graph convolutional network” This was an adaptation of the search methodology outlined in a similar scoping review by Chatterjee *et al*. (16) The search strategy was agreed upon by the authors (SB, JZ, HA, NS).

### Inclusion and exclusion criteria

Publications and preprints available until 21 November 2021 written in English were included. Manuscripts were required to (i) describe the creation of a KG or the use a KG to generate insights; (ii) use multimodal graphs (i.e. graphs with more than one discrete node class); and (iii) directly mention medical insights and/or health outcomes in humans.

We excluded (i) neuroimaging graph theory and graph topology manuscripts, unless KGs were explicitly mentioned within the manuscript;(ii) papers analysing animal models; (iii) articles about traffic road safety where health outcomes were not explicitly mentioned; (iv) manuscripts using graph neural network or graph theory which did not make use of a KG; (v) reviews and commentary articles, opinion pieces, editorials, and any other articles that did not report original research; (vi) single modality graphs with only one node class (as defined by the authors); and (vii) papers not in English language. Review contents were used to guide additional literature discovery.

### Data Collection

Duplicate manuscripts were removed using Endnote and Rayyan.ai. Two reviewers (JZ, SB) carried out a title and abstract screening. Papers passing the abstract screen underwent a full-text screening by two reviewers (SB and one of JZ, YA, VS, DM, EF, and MA). Data extraction was carried out by two reviewers (SB and one of JZ, YA, VS, DM, EF, and MA).

For manuscripts passing full-text screen, we collected the following data:

i. Demographics: (1) domain (a preliminary categorisation was refined iteratively by SB and JZ, with final categorization consisted of: medical science insights, drug repurposing, literature representation, drug interactions and toxicity, diagnostics, drug discovery, electronic health record (EHR) data representation, public health, non-EHR patient data representation, risk prediction, and drug related uses outside of discovery, repurposing, interactions and toxicity) (2) affiliation of authors (institution and country) (3) disease-area (e.g. refined iteratively categorized by SB and JZ, final categorisation included: general/non-disease specific, infectious diseases (Covid-19, cancer, neurology, mental health, diabetes/endocrine, rare disease, respiratory disease)
ii. Graph characteristics and data sources: (1) public availability of the KG (2) node and edge types (e.g. genes, drugs, proteins, etc.) (3) named data sources (e.g. The Cancer Genome Atlas, MEDLINE scraping, private EHR data, etc.) (4) size of the graph (the number of nodes, edges, node classes and edge classes, if included in the manuscript)
iii. Analysis: (1) analysis methodology (see categorisation of analysis methodology in Supplementary Methods section for further details) (2) Validation: whether there was inside graph or outside graph (downstream) validation of insights (3) Future plans/next steps in the analysis

Descriptions of the categories for use case and analysis methodology are summarized in the Supplementary Methods section.

Regarding validation: inside graph validation refers to efforts to validate findings by dividing data into training, testing and validation datasets, by holding out or adding in datasets to check robustness of conclusions, using cross-validation techniques when running models, or using multiple analysis methods and comparison of performance scores. Outside graph validation refers to additional analyses to test findings generated from the original graph, which may include testing hypotheses using in vitro studies, animal experiments, human observational data, or clinical trials.

### Article filtering

Articles were filtered through abstract and full-text screen, as shown by the flow chart in Figure 1.

**Figure 1.**
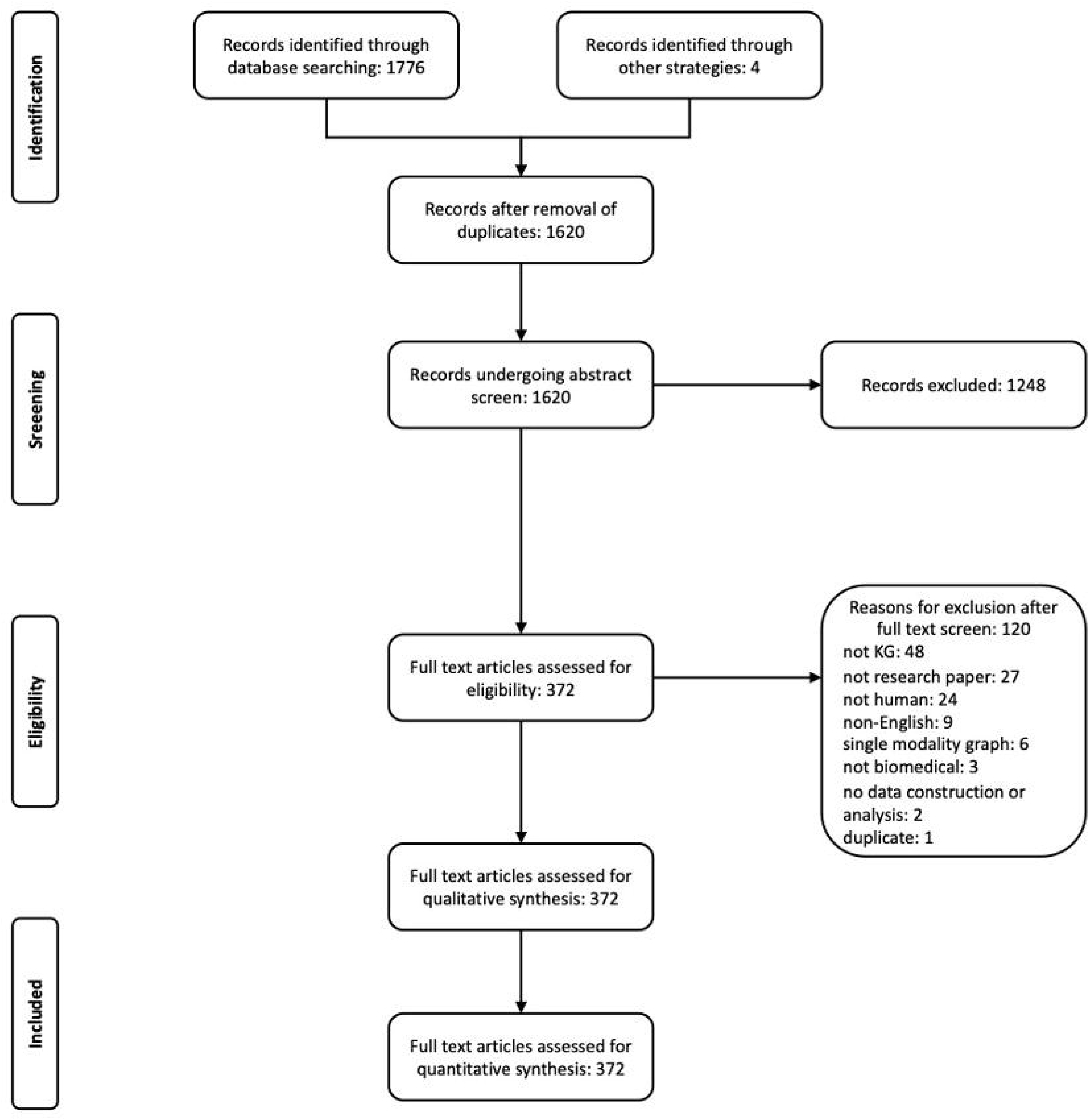

## Results

This section first presents demographic features of the surveyed manuscripts, including a description of manuscript numbers over time, use case, and disease area categorisation, author affiliations and funding sources. The graph characteristics session provides descriptive statistics between node and edge numbers, and node and edge class numbers in graphs used by the articles. A meta-graph of node classes is used to provide further information on node class characteristics from included graphs. A summary of data sources used in graph construction is presented. Finally, graph analysis techniques, validation methods and stated plans for future work are summarised.

### Manuscript demographics

Figure 2 demonstrates the trend in the number of manuscripts that were preprinted/published each year. There is an increasing number of publications in this area; furthermore, the trend has accelerated in recent years.

**Figure 2.**
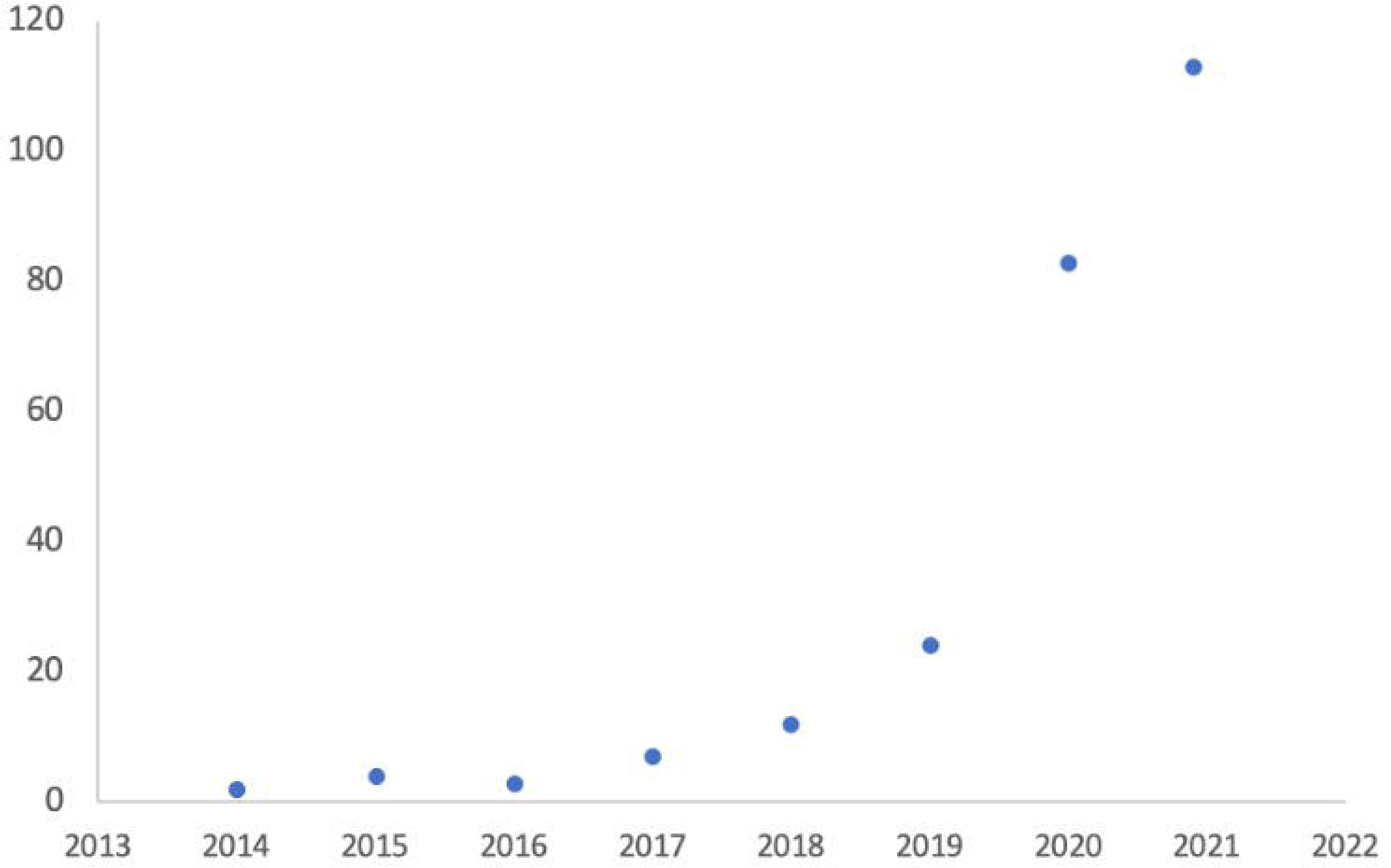

Figure 3 represents the categories of biomedical and healthcare knowledge graphs identified, grouped by use case. The two most common use cases are medical science insights unrelated to drugs or diagnostics (74 studies) and drug repurposing (57 studies), which are both twice as frequently seen as the third most common use case of literature representation (25). Papers categorised in the use case ‘medical sciences insights’ develop predictions or hypotheses regarding scientific knowledge. Examples might include protein-protein interactions, genes implicated in diseases, or a clustering of symptoms. Papers categorised as ‘literature representation’ include mapping and representation of research literature. This category includes bibliometrics and excludes papers which would be better categorised under other use cases such as diagnostics, drug repurposing, or other drug-related use cases. There is a broad array of use cases which include biomedical (medical science insights, drug repurposing, literature representations, drug interactions and toxicity, drug discovery, and drug related-other), patient related (EHR representation, non-EHR patient data) and population health management (diagnostics, public health, risk prediction). Descriptions of the use case categories are summarized in the Supplementary Methods section.

**Figure 3.**
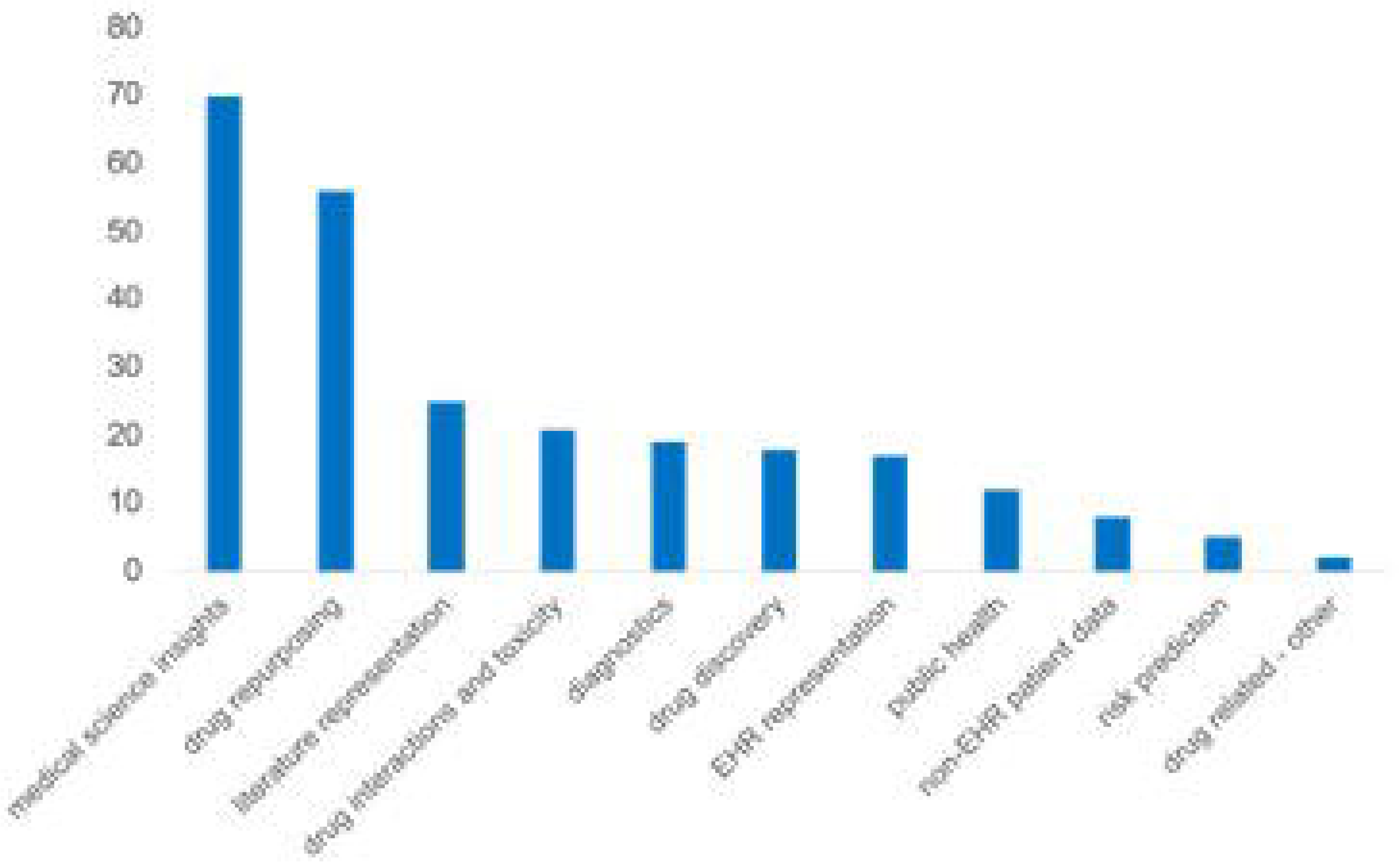

Figure 4 demonstrates the therapeutic area categorisation of identified manuscripts. General KGs are the most common category (115 manuscripts) and exceed KGs representing any individual therapeutic area. The most common therapeutic area was infectious disease (47 manuscripts); however, this was driven mainly by COVID-19 (41 manuscripts); KG for other infectious diseases (6 manuscripts) were less common. Outside of infectious diseases, manuscripts using KGs for oncology (26 manuscripts) and neurology (13 manuscripts) were also common. Graphs were often constructed to represent a single disease rather than a whole disease area.

**Figure 4.**
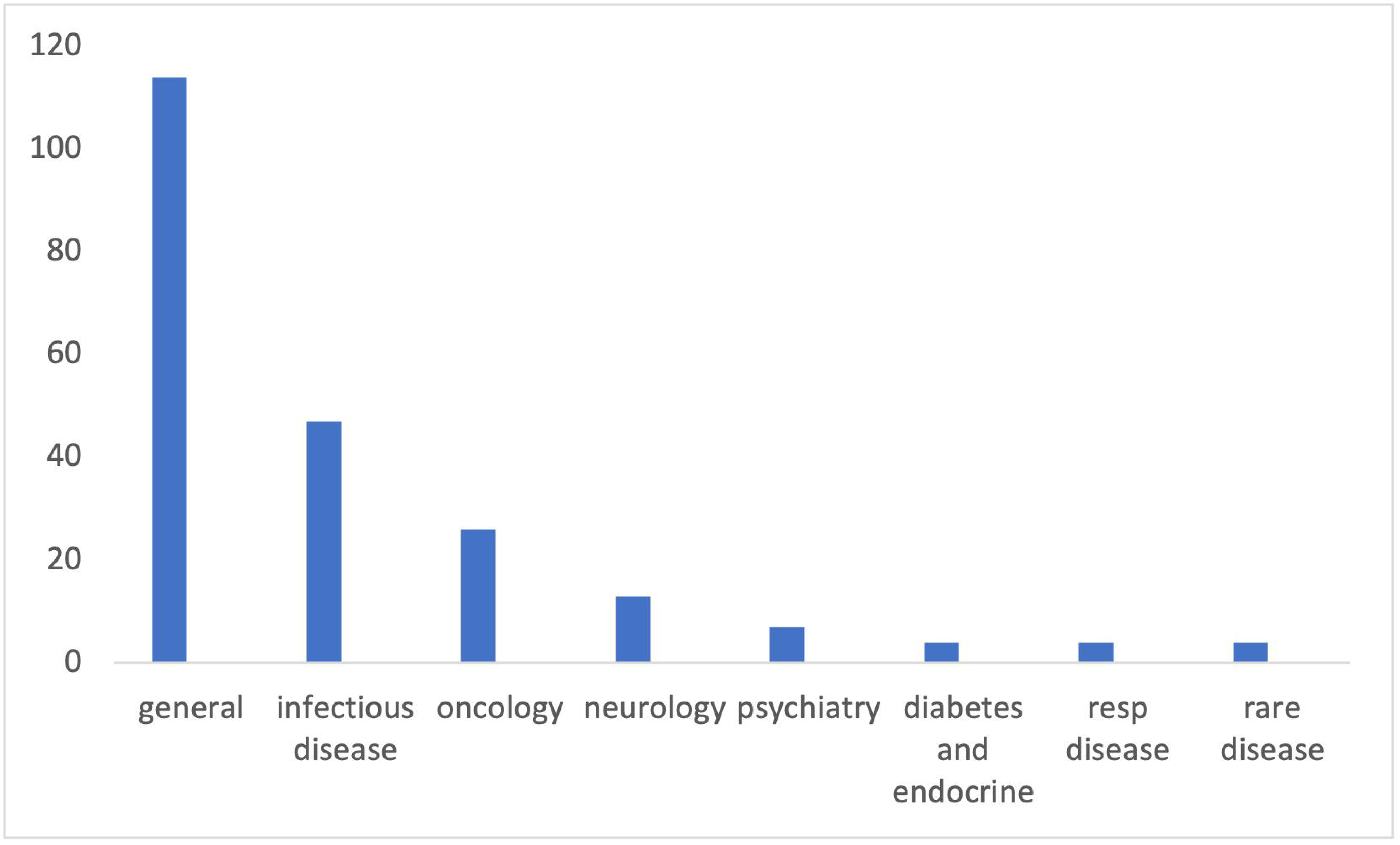

Table 1 demonstrates an aggregated count of country of primary affiliation for first and last authors of manuscripts. The United States and China are the top two contributors, with 162 and 155 manuscripts, respectively, followed by other European and North American countries. After USA and China, the next highest author count is from the UK (33 authors).

**Table 1.**

| Country | Count |
| --- | --- |
| USA | 162 |
| China | 155 |
| UK | 33 |
| Germany | 24 |
| Canada | 20 |
| Netherlands | 15 |
| Italy | 12 |
| Rest of Europe | 33 |

Table 2 demonstrates the breakdown of funding declarations within the manuscripts passing review screening. Each manuscript can have more than one archetype of funding source. Most manuscripts in this area receive funding for research from government or government funded bodies. More manuscripts receive commercial funding than non-governmental non-profit funding in this area. Most papers don’t have commercial affiliations. Tables 3 outlines the breakdown of companies funding KG research. There is not one single dominant company in this area. Multiple different company types have funded research in this area, including big technology companies, biopharmaceutical companies, consultancies and start-ups, with no single sector dominant.

**Table 2.**
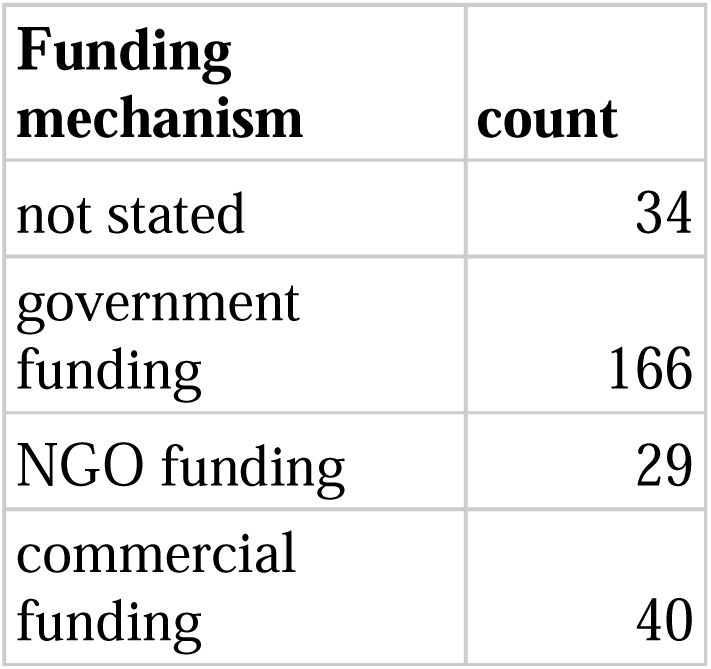

**Table 3.**

| **Company** | **Count** |
| --- | --- |
| Amazon (including AWS) | 5 |
| AstraZeneca | 5 |
| IBM | 5 |
| Google | 4 |
| Benevolent AI | 3 |
| CoVar Applied Technologies | 3 |
| Elsevier | 3 |
| Enveda Biosciences | 3 |
| Bayer | 2 |
| Biogen | 2 |
| Causality Biomodels | 2 |
| Data2Discovery | 2 |
| Euretos | 2 |
| IQVIA | 2 |
| nference | 2 |
| QIAGEN | 2 |
| Yidu Cloud | 2 |

### Graph characteristics

38.9% of manuscripts featured graphs that were open sourced. Our definition of open source is that the graph was downloadable or otherwise freely and openly available.

Table 4 provides summary statistics for unique node and edge classes within each graph. More manuscripts reported node classes than edge classes. In order to standardize the treatment of data across manuscripts reporting edge classes in a heterogenous manner, all edges between two node classes were counted as a single edge class. The skew and kurtosis of node and edge class counts suggest non-normally distributed data for both, so the median interquartile range (IQR) and range are presented. There is a wide range of node class numbers (2-41), but the median and IQR suggest this is due to outlying values. The mean class number is 6.0 and median is 4. The mean edge class number is 8.0, and the median is 4. The IQR for edge class numbers is 3 to 7. The range spans from a minimum of 1 to a maximum of 210, influenced by outlying values.

**Table 4.**

| **Value** | **Node class numbers** | **Edge class numbers** |
| --- | --- | --- |
| Number of values | 241 | 207 |
| Skewness | 3.3 | 9.3 |
| kurtosis | 15.0 | 109.5 |
| Mean | 6.0 | 8.0 |
| Median | 4 | 4 |
| Lower IQR | 3 | 3 |
| Upper IQR | 7 | 7 |
| Lower bound | 2 | 1 |
| Upper bound | 41 | 210 |

Table 5 summarises node and edge numbers reported by manuscripts. Fewer manuscripts report node and edge numbers compared to node class and edge class numbers. For node and edge numbers, the skewness and kurtosis of the datasets together suggest non-normal distribution, so median IQR and range are presented. The node number dataset includes 123 reports, and the edge numbers include 115 reports. The mean number of nodes across in KGs used by manuscripts is approximately 3 016 830, while the median is 46 983. The node number IQR is 454 533 but the range is higher at 180 199 788, suggesting outlier values. The mean number of edges across the KGs is 152 556 781, higher than the median which is 906 737, demonstrating skewed distribution. The edge number upper IQR is 9 828 637 but the range is higher at 13 999 999 762, due to outlier values. The ratio of median edge number to median node number was approximately 19, which gives some indication of the connectivity within graphs.

**Table 5.**

| **Value** | **Node numbers** | **Edge numbers** |
| --- | --- | --- |
| Number of values | 123 | 115 |
| Skewness | 9.6 | 10.6 |
| kurtosis | 99.5 | 112.8 |
| mean | 3 016 830 | 152 556 781 |
| median | 46 983 | 906 737 |
| lower IQR | 6 415 | 66 272 |
| upper IQR | 460 948 | 9 894 909 |
| lower bound | 212 | 238 |
| upper bound | 180 200 000 | 14 000 000 000 |

Figure 5 summarizes information about common node classes and their relations into a meta-graph. The nodes are aggregated node class categories extracted from the manuscripts, and edges represent the node class-node class relationships that are extracted from the manuscript. The methods for generation node class groupings and graph visualisation are further detailed in the Supplementary Methods section. They demonstrate that disease-gene and disease-drug edges are most commonly seen. On visual inspection, the graph separates into two groups centred upon disease concepts: biomedical nodes (here used to mean concepts more related to molecular biology, ‘wet lab’ science or bioinformatics) and clinical nodes (here used to mean concepts predominantly patient related or more commonly encountered in the patient facing context).

**Figure 5.**
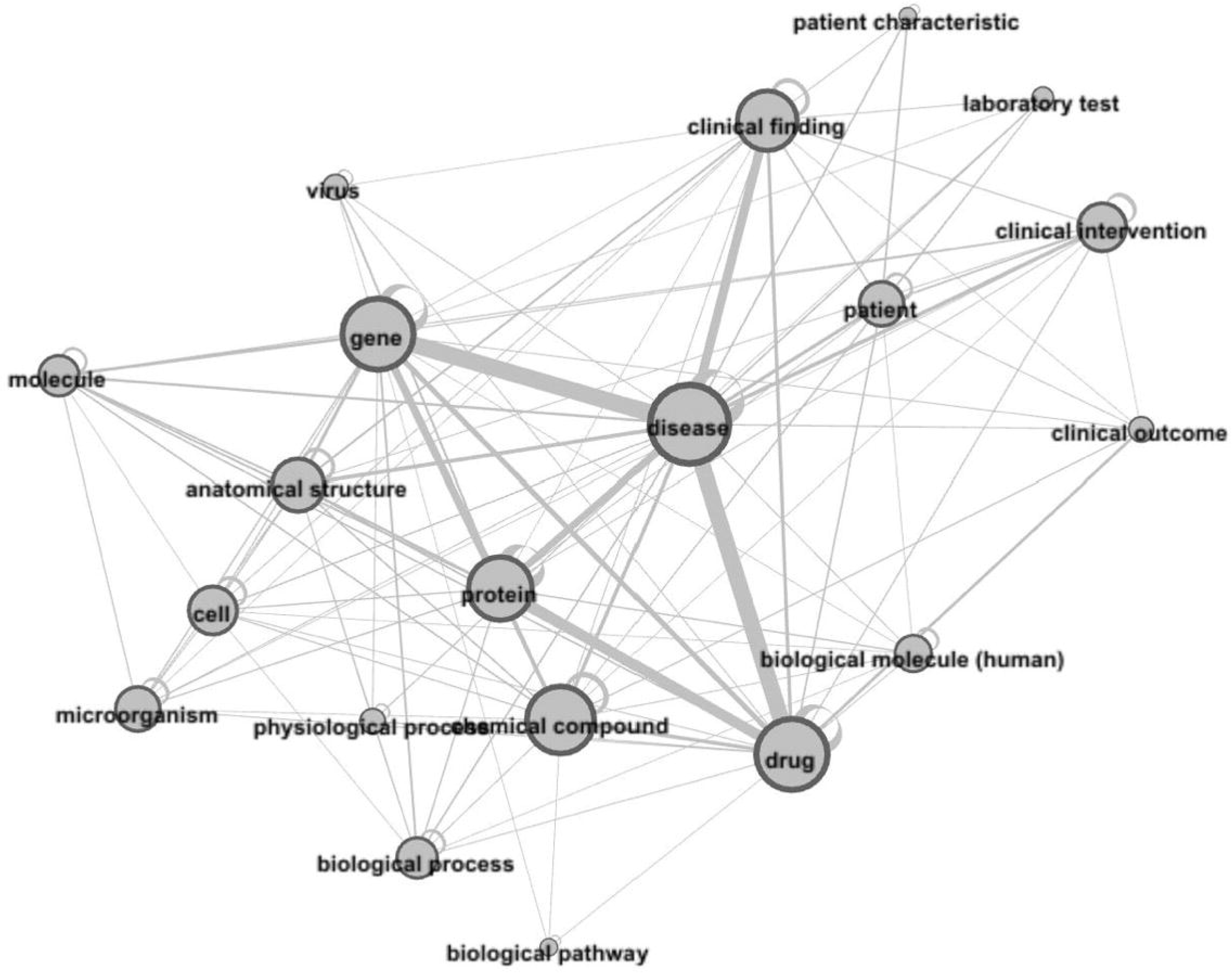

### Data sources

Table 6 provides a list of all datasets where five or more manuscripts used the source. DrugBank emerges as a frequently used data source, with utilization in 46 manuscripts, underscoring its significance in drug-related KG research. PubMed and UniProt also exhibit high usage, appearing in 26 KGs each, reflecting the importance of literature and protein data.

**Table 6.**
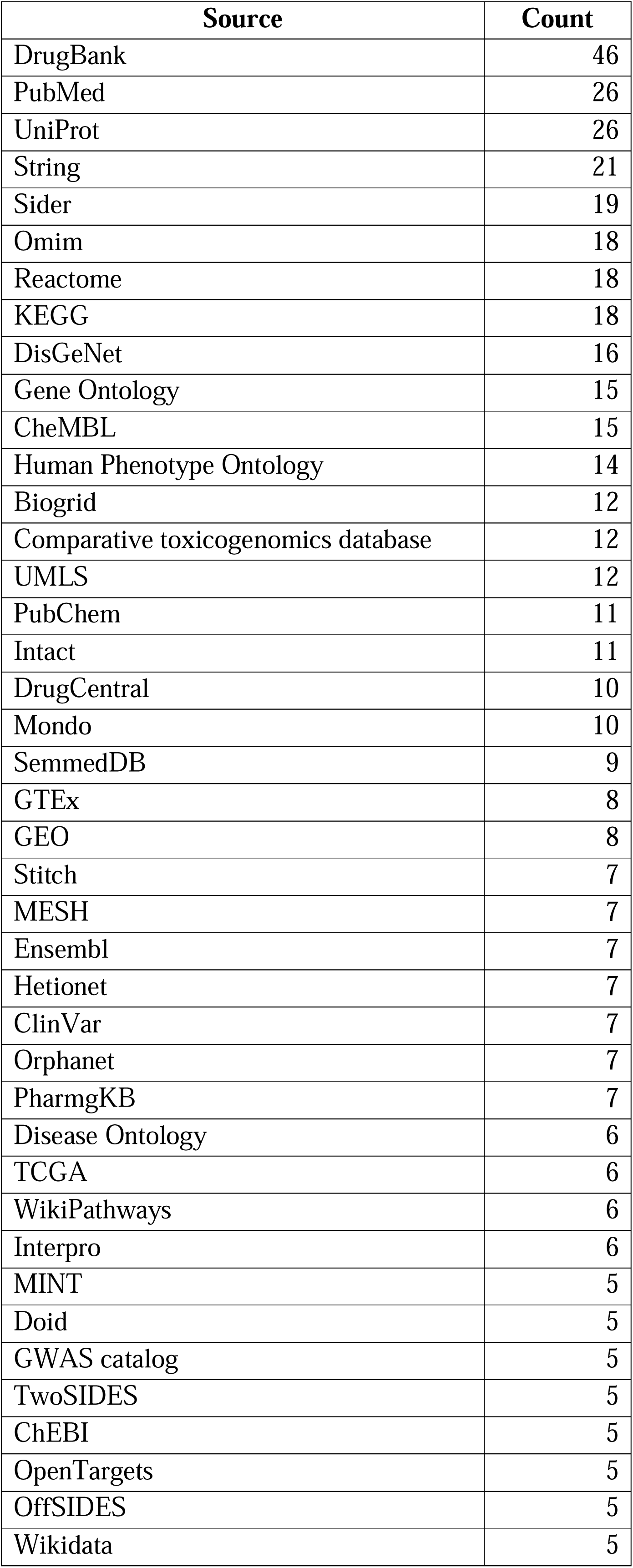
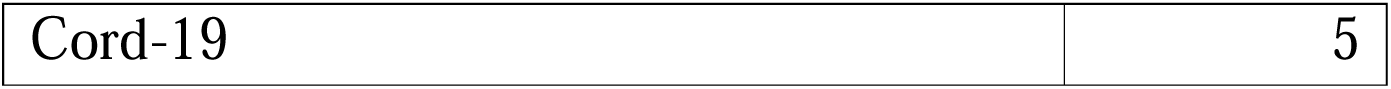

### Graph analysis

Table 7 summarizes counts of analysis technique archetypes used in KG analysis in the identified manuscripts. There are many different methodologies being used in KG analysis, of varying sophistication. We observed a diverse range of approaches in KG research. The most prevalent method is graph querying, used in 52 instances, followed by graph embedding (43) and Graph Convolutional Networks (GCNs) (33).

**Table 7.**

| **Analysis method breakdown** | **Count** |
| --- | --- |
| querying | 52 |
| graph statistics | 24 |
| node embedding | 20 |
| graph embedding | 43 |
| supervised classification methods (non-embedding) | 17 |
| unsupervised graph clustering | 13 |
| GCN | 33 |
| graph attention networks | 14 |
| deep learning (other) | 17 |
| other | 14 |

Table 8 presents the breakdown of validation methods employed in the identified manuscripts. Most papers (120) focus on inside graph validation techniques, such as data splitting, cross-validation, and algorithm diversity to ensure robustness. Less than a third of papers (39) validate findings outside of the graph. Such validation techniques may include *in vitro* testing, or clinical trials. 37 manuscripts combine within and outside graph validation approaches.

**Table 8.**
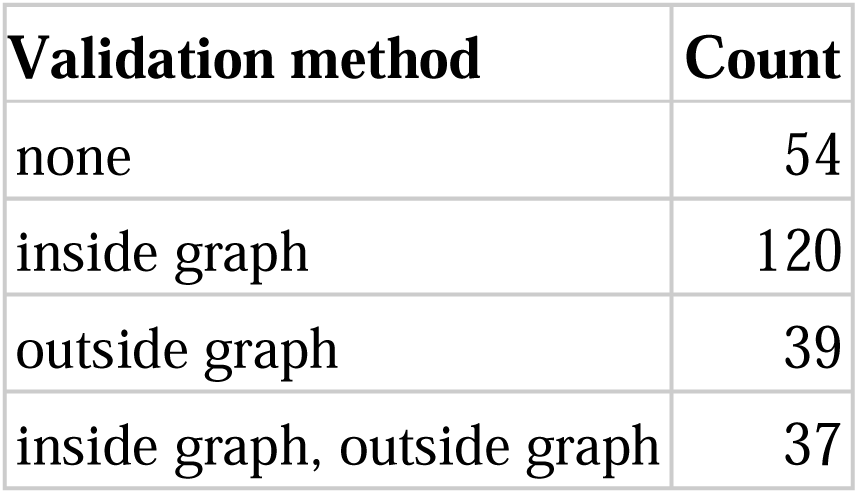

Table 9 provides a summary of planned or suggested next steps that authors have stated in Discussion sections of their manuscript The most frequently discussed future directions are data and algorithms, with 99 and 59 counts. Categories aligned with validation (clinical trials, other validation) feature less frequently at 3 and 11 counts.

**Table 9.**

| **Future plan category** | **Count** |
| --- | --- |
| improve data | 99 |
| improve algorithm | 59 |
| extend use case | 42 |
| other validation | 21 |
| improve user interface | 18 |
| clinical application | 11 |
| clinical trials | 3 |
| none | 25 |

Table 10 provides a summary of Tables and Figures as related to aims of the scoping review, as recommended by PRISMA-ScR guidelines. (18)

**Table 10.**

| **Aim of scoping review** | **Relevant Tables and Figures** |
| --- | --- |
| use-cases | Figures 3, 4 |
| data characteristics | Table 4, 5, 6; Figure 5 |
| research characteristics | Tables 1, 2, 3, 7, 9; Figure 2 |
| validation | Table 8 |

## Discussion

### Findings and implications

KGs are becoming increasingly used in biomedical and healthcare sciences. The most prevalent applications of KGs are in medical science insights and drug repurposing – this is reflected in the common use of drug and protein datasets in KG construction. Research activity is concentrated in North America, China, and Europe. Location of research, together with location of data sources, contributes to bias in dataset curation for openly available datasets. (19) Despite significant commercial interest, the majority of academically published research in this domain continues to be government funded.

The variation in KG utilization suggests that there are potential opportunities in use cases and disease/therapeutic areas where knowledge graphs have seen limited use. KGs tend to either focus on specific diseases or remain highly general, with fewer encompassing entire therapeutic areas or multiple domains. A challenge arises from enrichment of KGs with data for a specific disease; this approach may improve predictions when analysing a disease-relevant question by adding relevant contextual data, but may introduce bias into the graph.(20) (21)

KGs exhibit substantial heterogeneity in terms of size, as reflected by wide variation in node and edge counts, as well as counts of node and edge classes. This may in part reflect the diverse scope and use cases for knowledge graphs or represent limitations in available storage and compute for analysis but may also represent a lack of known best practice regarding optimal KG size and connectivity.

This study reveals a wide array of data sources used in KG construction, with a preference for open datasets over closed ones. Despite the diversity, there is a clear concentration with some datasets, for example DrugBank, used frequently. Even the ostensible diversity hides a network of dependencies where certain datasets rely on others for primary data sources. For example, Open Targets uses many sources, including UniProt, STRING, Reactome, and ChEMBL. (22) More generally, there are biases stemming from input data. For example, there is a correlation between information available for bio-entities and NIH funding for research into those entities. (23)

Figure 5 suggests that on visual inspection, there is a grouping of biomedical node classes (here taken to mean concepts more often found in molecular biology or other ‘wet lab’ science or bioinformatics) and a grouping of clinical node classes (here used to indicate concepts predominantly patient-related or more commonly encountered in the patient facing context).

This grouping of biomedical and clinical node classes may be due to different data pipelines that would be used to generate node classes in each cluster and the relative ease of capturing some concepts and edges in one type of graph versus the other.

Biomedical node classes often use curated datasets where data has been methodically organized, corrected, annotated, and standardized through mapping and harmonizing to various ontologies and vocabularies. (24) This data is more likely to be available in the public domain through publications, hence the denser connectivity in this cluster.

In contrast, the construction of graphs using clinical node classes is more challenging. (25) First, EHR and public data backends are more individualised. For example, different hospitals may use different coding ontologies, electronic health records, and file storage types and methods from those systems. This lack of common standards makes transfer between health applications challenging. Second, healthcare data tends to be noisy, with large amounts of missingness and censoring, including loss of follow-up, incomplete data recording from patients who present to a care setting, and challenges in capturing phenomic data such as lifestyle and diet. This results in systematic biases in data. Third, there is a data structuring challenge. EHRs contain large amounts of unstructured text, which requires processing and may need either additional, highly expensive manual curation or improved automated curation, for example, through large language models. (26)(27) Non-text data such as radiologic and pathologic images would require transformation for ingestion to preserve underlying, complex features and latent variables that might be lost in simple feature extraction. Fourth, regulation may require higher privacy safeguards to ensure that released data does not contain patient details. In the USA, this is governed through the Health Insurance Portability and Accountability Act (HIPAA).

Aside from comparisons between graphs predominantly using biomedical or healthcare node classes, the breadth of use cases depends on graph design, which involves dataset inclusion, schema flexibility, and expansiveness of relevant vocabularies and ontologies. Graphs may also have broad coverage of diseases or be a narrower representation of a single disease or disease group. Though it is likely from current evidence that graph performance will scale in size, performance has also been demonstrated to improve with context. (28) This means that it is likely no single best all-purpose graph. Instead, there is a judgement based on the graph use-case.

The prevalence of graph querying in KG research is surprising given the simplicity of the approach. However, this may be appropriate to the research objectives of those manuscripts if this is for data exploration or transparency in insight generation. Graph machine learning (ML) can perform a wider variety of tasks, for example, link prediction, node classification, or community detection. Graph ML can help mitigate data biases, for example, through graph rewiring or regularization techniques.

Few manuscripts include outside graph validation (Table 8) or explicitly suggest outside graph validation as further work (Table 9). This is concerning but understandable. There may be constraints on carrying out *in vitro* or *in vivo* experimental work or trials due to budgetary restrictions, regulatory constraints, lack of domain expertise, or challenges coordinating cross-disciplinary work. Outside graph validation would help translate insights from KGs into real-world applications.

## Future work

There are several areas for further research. First, best practice in KG construction, especially concerning graph size, remains unresolved. Future reviews should further analyze specific use cases of KGs to interrogate tailored outcome measures for those graphs. Identifying factors associated with successful outcomes could inform best graph construction and analysis practices. In addition, it would be helpful to compare the effectiveness of KGs against alternative techniques for data source integration. Second, investigating ways to enhance the integration of -omics data and patient data within KGs could advance personalized medicine and disease understanding. Third, as the paper selection for this review concluded before the rising prevalence of generative AI and large language models (LLMs), an exciting next phase in the field involves understanding how to integrate KGs with these techniques, presenting new possibilities for knowledge representation and utilization. (29) (30) Advancements in LLMs will facilitate the incorporation of unstructured data sources such as EHR data. Fourth, given the low percentage of graphs that are open sourced to consider how best to encourage sharing and coordination of KGs in this domain. An example is the Therapeutic Data Commons initiative. (31) One barrier to sharing graphs using clinical data may be regulatory and data privacy concerns.

Ultimately, insights from KGs have yet to realise their potential in delivering clinically actionable findings, especially in patient-facing settings. Adding clinical data to biomedical graphs may be beneficial for drug repurposing and generation of medical science insights. Clinical data itself may be used to better understand disease clustering, patient pathways, and in construction of digital twins. In order to improve the use of clinical data for combination with biomedical data or on its own, the challenges are technical (ontology standardisation, data ingestion) and regulatory (ensuring de-identification, and legal clearance for data sharing). To overcome the challenge of using proprietary data sources, it is possible to use data that can be accessed on applications, such as MIMIC (32) or use publicly released graphs (25).

## Limitations

This study has limitations, including the November 2021 screen cut-off date for manuscript inclusion, which may have excluded more recent developments in KGs. The criteria used for manuscript selection may have inadvertently excluded relevant studies, for example, those that use graphs but do not include this detail in keywords or MESH terms. As there is commercial activity in this area, complementary datasets such as patent screens might provide additional insights into the use of KGs in this area. In conducting this initial scoping review, we established categorizations for use cases, disease areas, and analysis methods through a consensus-driven process involving authors SB and JZ with input from HA and NS. This was followed by an iterative refinement during review, based primarily on ease of manuscript categorisation and identification of areas where manuscripts did not clearly belong to a category. These categorizations are a function of the exploratory nature of this study. Future research may further refine and adapt this framework of categorizations.

## Conclusion

In summary, KGs have many possible uses within in biomedicine and healthcare, but their full potential is yet to be realised. The two most popular use cases to date are generation of medical science insights and drug repurposing. There is an opportunity to expand work areas across other use cases and across diseases. Heterogeneity in graph size and context specificity suggests further work is needed to understand optimum graph construction. There are many different techniques used in graph analysis - deploying more sophisticated graph machine learning techniques may improve insights gained from KGs. Validation of findings from graphs through external testing will increase the robustness of conclusions drawn from graphs. While there are many graph-specific factors preventing realisation of utility of KGs in clinical settings, there are many more general barriers to implementation in clinical AI. (33) (30)

## Data Availability

All data produced in the present study are available upon reasonable request to the authors

## References

1. Zhang J, Symons J, Agapow P, Teo JT, Paxton CA, Abdi J, et al. Best practices in the real-world data life cycle. PLOS Digital Health. 2022 Jan 18;1(1):e0000003.

2. Krassowski M, Das V, Sahu SK, Misra BB. State of the Field in Multi-Omics Research: From Computational Needs to Data Mining and Sharing. Frontiers in Genetics [Internet]. 2020 [cited 2023 Nov 27];11. Available from: https://www.frontiersin.org/articles/10.3389/fgene.2020.610798

3. Ji S, Pan S, Cambria E, Marttinen P, Yu PS. A Survey on Knowledge Graphs: Representation, Acquisition, and Applications. IEEE Transactions on Neural Networks and Learning Systems. 2022 Feb;33(2):494–514.

4. Peng C, Xia F, Naseriparsa M, Osborne F. Knowledge Graphs: Opportunities and Challenges. Artif Intell Rev. 2023 Nov 1;56(11):13071–102.

5. Patel VL, Evans DA, Groen GJ. Biomedical knowledge and clinical reasoning. 1989;

6. Kumar K, Palakal MJ, Mukhopadhyay S, Stephens MJ, Li H. BioMap: toward the development of a knowledge base of biomedical literature. In: Proceedings of the 2004 ACM symposium on Applied computing [Internet]. New York, NY, USA: Association for Computing Machinery; 2004 [cited 2023 Nov 17]. p. 121–7. (SAC ’04). Available from: https://dl.acm.org/doi/10.1145/967900.967927

7. Geleta D, Nikolov A, Edwards G, Gogleva A, Jackson R, Jansson E, et al. Biological Insights Knowledge Graph: an integrated knowledge graph to support drug development [Internet]. bioRxiv; 2021 [cited 2022 Nov 17]. p. 2021.10.28.466262. Available from: https://www.biorxiv.org/content/10.1101/2021.10.28.466262v1

8. Smith DP, Oechsle O, Rawling MJ, Savory E, Lacoste AMB, Richardson PJ. Expert-Augmented Computational Drug Repurposing Identified Baricitinib as a Treatment for COVID-19. Frontiers in Pharmacology [Internet]. 2021 [cited 2022 Sep 25];12. Available from: https://www.frontiersin.org/articles/10.3389/fphar.2021.709856

9. Chen X, Jia S, Xiang Y. A review: Knowledge reasoning over knowledge graph. Expert Systems with Applications. 2020 Mar 1;141:112948.

10. Zamini M, Reza H, Rabiei M. A Review of Knowledge Graph Completion. Information. 2022 Aug;13(8):396.

11. Nickel M, Murphy K, Tresp V, Gabrilovich E. A Review of Relational Machine Learning for Knowledge Graphs. Proceedings of the IEEE. 2016 Jan;104(1):11–33.

12. Issa S, Adekunle O, Hamdi F, Cherfi SSS, Dumontier M, Zaveri A. Knowledge Graph Completeness: A Systematic Literature Review. IEEE Access. 2021;9:31322–39.

13. Ye H, Zhang N, Chen H, Chen H. Generative Knowledge Graph Construction: A Review [Internet]. arXiv; 2023 [cited 2023 Oct 3]. Available from: http://arxiv.org/abs/2210.12714

14. Zeng X, Tu X, Liu Y, Fu X, Su Y. Toward better drug discovery with knowledge graph. Current Opinion in Structural Biology. 2022 Feb 1;72:114–26.

15. Nicholson DN, Greene CS. Constructing knowledge graphs and their biomedical applications. Computational and Structural Biotechnology Journal. 2020 Jan 1;18:1414– 28.

16. Chatterjee A, Nardi C, Oberije C, Lambin P. Knowledge Graphs for COVID-19: An Exploratory Review of the Current Landscape. Journal of Personalized Medicine. 2021 Apr;11(4):300.

17. Li MM, Huang K, Zitnik M. Graph representation learning in biomedicine and healthcare. Nat Biomed Eng. 2022 Dec;6(12):1353–69.

18. Tricco AC, Lillie E, Zarin W, O’Brien KK, Colquhoun H, Levac D, et al. PRISMA Extension for Scoping Reviews (PRISMA-ScR): Checklist and Explanation. Ann Intern Med. 2018 Oct 2;169(7):467–73.

19. Futoma J, Simons M, Panch T, Doshi-Velez F, Celi LA. The myth of generalisability in clinical research and machine learning in health care. The Lancet Digital Health. 2020 Sep 1;2(9):e489–92.

20. Pujara J, Augustine E, Getoor L. Sparsity and Noise: Where Knowledge Graph Embeddings Fall Short. In: Proceedings of the 2017 Conference on Empirical Methods in Natural Language Processing [Internet]. Copenhagen, Denmark: Association for Computational Linguistics; 2017 [cited 2023 Oct 8]. p. 1751–6. Available from: https://aclanthology.org/D17-1184

21. Mohamed A, Parambath S, Kaoudi Z, Aboulnaga A. Popularity Agnostic Evaluation of Knowledge Graph Embeddings. In: Proceedings of the 36th Conference on Uncertainty in Artificial Intelligence (UAI) [Internet]. PMLR; 2020 [cited 2023 Oct 8]. p. 1059–68. Available from: https://proceedings.mlr.press/v124/mohamed20a.html

22. Ochoa D, Hercules A, Carmona M, Suveges D, Baker J, Malangone C, et al. The next-generation Open Targets Platform: reimagined, redesigned, rebuilt. Nucleic Acids Research. 2023 Jan 6;51(D1):D1353–9.

23. Tan F, Zhang T, Yang S, Wu X, Xu J. Discovering Booming Bio-entities and Their Relationship with Funds. Data and Information Management. 2021 Jul 1;5(3):312–28.

24. Orchard S, Hermjakob H. Shared resources, shared costs—leveraging biocuration resources. Database. 2015 Jan 1;2015:bav009.

25. Hong C, Rush E, Liu M, Zhou D, Sun J, Sonabend A, et al. Clinical knowledge extraction via sparse embedding regression (KESER) with multi-center large scale electronic health record data. npj Digit Med. 2021 Oct 27;4(1):1–11.

26. Yang X, Chen A, PourNejatian N, Shin HC, Smith KE, Parisien C, et al. A large language model for electronic health records. npj Digit Med. 2022 Dec 26;5(1):1–9.

27. Bean DM, Kraljevic Z, Shek A, Teo J, Dobson RJB. Hospital-wide natural language processing summarising the health data of 1 million patients. PLOS Digital Health. 2023 May 9;2(5):e0000218.

28. Li MM, Huang Y, Sumathipala M, Liang MQ, Valdeolivas A, Ananthakrishnan AN, et al. Contextualizing protein representations using deep learning on protein networks and single-cell data [Internet]. bioRxiv; 2023 [cited 2023 Nov 20]. p. 2023.07.18.549602. Available from: https://www.biorxiv.org/content/10.1101/2023.07.18.549602v1

29. Zhao J, Zhuo L, Shen Y, Qu M, Liu K, Bronstein M, et al. arXiv.org. 2023 [cited 2023 Oct 8]. GraphText: Graph Reasoning in Text Space. Available from: https://arxiv.org/abs/2310.01089v1

30. Pan JZ, Razniewski S, Kalo JC, Singhania S, Chen J, Dietze S, et al. arXiv.org. 2023 [cited 2023 Oct 8]. Large Language Models and Knowledge Graphs: Opportunities and Challenges. Available from: https://arxiv.org/abs/2308.06374v1

31. Huang K, Fu T, Gao W, Zhao Y, Roohani Y, Leskovec J, et al. Artificial intelligence foundation for therapeutic science. Nat Chem Biol. 2022 Oct;18(10):1033–6.

32. Johnson AEW, Bulgarelli L, Shen L, Gayles A, Shammout A, Horng S, et al. MIMIC-IV, a freely accessible electronic health record dataset. Sci Data. 2023 Jan 3;10(1):1.

33. Zhang J, Budhdeo S, William W, Cerrato P, Shuaib H, Sood H, et al. Moving towards vertically integrated artificial intelligence development. npj Digit Med. 2022 Sep 15;5(1):1–9.

34. Inc S. fuzzywuzzy: Fuzzy String Matching in Python. [Internet]. 2014. Available from: https://github.com/seatgeek/fuzzywuzzy.

