## Supplementary Methods for "Scoping review of knowledge graph applications in biomedical and healthcare sciences"

**Data collection**

***Title, abstract and full text screens***

Discrepancies in the title an abstract screen were reviewed by JZ and SB after unblinding, to reach a consensus decision. For discrepancies in the full text screen, a third reviewer (JZ or YA) reviewed the manuscript.

***Categorisation in data collection***

Preliminary categorisations were defined *a priori* and refined during data collection

***Categorisations of use case***

The following definitions were used as a guide for final use case categorisations:

- medical science insights: Predictions or hypotheses regarding scientific knowledge. Examples might include protein-protein interactions, genes implicated in diseases, or a clustering of symptoms
- drug repurposing: identification of novel indications for existing drugs
- literature representation: mapping and representation of research literature. This category includes bibliometrics. This excludes papers which are better categorised under diagnostics, drug repurposing, or other drug related categories
- drug interactions and toxicity: prediction of drug side effects or adverse events
- diagnostics: use of knowledge graphs to predict patient diagnosis. This may be based on electronic health records or other data sources, such as patient data entry on a virtual platform
- drug discovery: identification of novel drugs which may have future uses as treatments
- EHR representation: Mapping and representation of clinical and/or operational data held in electronic health records. EHR representation specifically for diagnostics is included in the diagnostics category
- public health: Representation of concepts for the purpose of or regarding population health management. This includes social determinants of health, environmental factors impacting health, and infectious disease outbreaks
- non-EHR patient data: Mapping and representation of patient data where the source is not from electronic health records, and the purpose is not related to other categories, including drug related categories (drug repurposing, drug discovery, drug interactions and toxicity, drug related - other) or diagnostics category. This includes patient data entry in cellular/mobile or web portals and use of claims data.
- risk prediction: Mapping and representations of data in order to predict or otherwise better appreciate risk of acquiring a condition, or of disease prognosis.
- drug related - other: representation of drug data for a purpose other than drug repurposing, drug discovery or drug interactions and toxicity. This includes representation of drug-target data without downstream use predictions regarding repurposing, interactions, etc.

***Categorisation of analysis methodology***

The following definitions were used as a guide for determining whether which category of knowledge graph analysis was used in each manuscript. Where possible only one category was chosen – for example, if a novel analysis methodology was compared to pre-existing methods to demonstrate superiority, only the novel method was categorised.

Querying: The process of extracting data from a graph using a specific query language or API. The graph might be queried to extract a subgraph. Examples may include finding all the nodes that meet certain criteria or retrieving specific information about a node.

Graph statistics: Analysing properties of a graph, including measures including but not limited to centrality, degree distribution, and clustering coefficient.

Graph convolutional networks (GCN): A type of neural network that can be used to learn from graphs by aggregating information from a node's neighbors through message passing.

Graph attentional networks: A neural network that can be used to learn from graphs by selectively attending to important nodes or edges using attention mechanisms. This allows graph attentional networks to focus on the most relevant information for a given task.

Deep learning (non-GCN, non-GAN): Deep learning is machine learning using neural networks with multiple layers. This includes methods such as graph autoencoders, Graph Recurrent Neural Networks, Graph Generative Adversarial Networks, but excludes GCNs or GANs which have a separate model label.

Embedding methods without use of GCN, GAN or deep learning architectures:

Node embedding: The process of representing a node as a vector in a low-dimensional space (i.e. simplifying complex data about the node while preserving important information), which captures the node's characteristics and context within the graph. This can be done using a variety of methods, e.g. node2vec, DeepWalk, LINE, SDNE

Graph embedding: The process of representing a graph as a fixed-length vector (i.e. string of numbers) or matrix (i.e. 2D table of numbers) that captures its structural properties and relationships between nodes. e.g. RotatE, TransE

Supervised classification methods (non-embedding): A type of machine learning that can be used to classify graphs by using labeled data to train a model. This includes methods such as support vector machines (SVMs), decision trees, random forests, logistic regression, K-Nearest Neighbours, or naive Bayes. Supervised learning uses labelled data, with desired output types already known.

Unsupervised graph clustering: machine learning to group or cluster nodes or graphs based on their structural similarities, without the data having labels. This includes methods such as spectral clustering, hierarchical clustering, and k-means clustering.

**Data analysis**

Categorization of use case, disease area, analysis type and future plans were iterative and based on periodic review and agreement between SB and JZ. Analysis was carried out by SB and JZ. Graph demographics were analyzed in Google Sheets. Harmonisation of dataset terms took place in a semi-automated fashion with Python, Pandas, fuzzywuzzy (34) and manual review. Harmonisation of node terms for Figure 5 to create gold standard concepts for grouping took place in a semi-automated fashion using Python, BERT PubMed embedding and k-nearest neighbors classification with manual review of groupings. Meta-graph visualisation of co-occurring node terms was carried out with Gephi.

34.	Inc S. fuzzywuzzy: Fuzzy String Matching in Python. [Internet]. 2014. Available from: https://github.com/seatgeek/fuzzywuzzy.
